# Development of a village health worker care model to reduce cardiovascular risk in areas of armed conflict: Qualitative results from a participatory planning process in Myanmar

**DOI:** 10.1101/2025.11.24.25340927

**Authors:** Eindra Htoo, Parveen Parmar, Adam Richards, Tara Russell, Catherine Lee, Saw Sha Kler Law, Saw Kyaw Myint, Zay Yar Phyo Aung, Tom Traill

## Abstract

Cardiovascular disease (CVD) is the leading cause of death in Myanmar, but care models are lacking to deliver evidence-based treatments to reduce CVD risks in areas of armed conflict. Village health workers (VHWs) play a key role in bridging communities and clinics, but their role in CVD prevention and management in conflict-affected settings has not been clearly defined. This study aimed to identify the functions of VHWs that could improve access and quality of CVD care in conflict-affected areas of Karen State, Myanmar. We conducted 33 key informant interviews with patients, VHWs, clinic staff, ethnic health organization leaders, donors, and non-government organization health experts. A qualitative content analysis was carried out to explore barriers, facilitators, and potential leverage points for improving CVD care in conflict-affected settings. Participants reported that many people were unaware of CVD and its risks, and there were persistent gaps in CVD screening, medication adherence, and referral systems. Transportation costs and hazards, medicine shortages, and security risks made it difficult for patients to visit clinics or achieve continuity of care. Respondents believed VHWs could play an important role in improving CVD care by providing health education sessions, screening for high blood pressure and diabetes, delivering medications and supporting adherence, and facilitating referrals. Results showed expanding VHW roles could make CVD care more accessible, particularly where travel to clinics was unsafe or unaffordable. This study underscores the urgent need to develop and test community-based strategies to mitigate the growing burden of CVD in conflict-affected Eastern Myanmar. VHWs have the potential to address CVD care gaps. Targeted training and regular supervision of VHWs, coupled with structured systems for referral and follow-up, could enhance continuity of care, adherence to treatment, and community resilience to acute conflict events.

## Introduction

Cardiovascular disease (CVD) is the leading cause of death and disability in many fragile and conflict-affected states, including Myanmar [1,2]. However, the humanitarian system has neglected to invest in CVD diagnosis and treatment; one study estimated that non-communicable diseases (NCDs), which include CVD, received only 0.44% of overseas development aid for health targeted to internally displaced persons (IDPs). Coverage of interventions proven to reduce CVD risk is low, and evidence is lacking to support the effectiveness of care models to diagnose and treat hypertension, diabetes, and elevated CVD risk (with statins) in areas of active conflict [3–5].

Karen State, Myanmar, exemplifies this global pattern of underinvestment and low service coverage for CVD risk factors. A military coup in 2021 acutely exacerbated access to CVD services for over 260,000 IDPs [6] who, after six decades of chronic armed conflict, have come to rely on Ethnic Health Organizations (EHOs) for most health services [7,8]. Health financing is dependent on donors who have prioritized maternal child health and infectious diseases such as malaria and tuberculosis over NCDs. Resources for CVD risk factor management are limited, and coverage is low [9].

In 2022, the Karen Department of Health and Welfare (KDHW) was funded to conduct a three-phase study to develop and test in a randomized controlled trial a village health worker (VHW) care model to increase access to CVD services in remote rural areas of Karen State experiencing violence due to armed conflict. KDHW chose to develop a VHW-centered care model based on their successful experience with decentralized approaches to maternity care and malaria control, delivered by maternal health and village malaria workers [10–12]. By bringing services to beneficiaries instead of requiring beneficiaries to travel to services, decentralized models can reduce transportation costs and other structural barriers to care and promote health equity [13]. Multiple studies support the effectiveness of community health worker (CHW) care models to control hypertension and other CVD risk factors in low and middle-income countries (LMICs), but their potential contribution to CVD prevention and management in conflict-affected areas has not been extensively explored [14–22]. A global priority-setting exercise ranked developing and testing the effectiveness of CHW care models as the top priority for research on cardio-metabolic syndrome in humanitarian emergencies [23].

Here we present results of qualitative interviews that supported the Phase 1 objective of the study, which was to develop a comprehensive strategy to screen and treat individuals at risk of heart attacks and strokes, to be tested in later study phases. The qualitative aims were to explore the current status of CVD care in the region, illuminate the strengths and limitations of VHWs, and examine how they might contribute to a village-based care model that overcomes structural and behavioral barriers.

## Methods

### Ethics Statement

This study was deemed exempt by the University of Southern California Office of Human Subjects Research and received ethical approval from the regional Community Ethics Advisory Board, which is a community-led group that provides ethical guidance and oversight for research. Funding was provided by Research for Health in Humanitarian Crises (R2HC/Elrha #78698) and a Research Innovation Award from the Office of Research Excellence, Milken Institute School of Public Health, George Washington University (#09RIA042022). All key informant interviews (KIIs) provided informed verbal consent.

This study was guided by a participatory, iterative development process to capture the complexity of healthcare for CVD risk management in a conflict-affected community health system, reflecting the perspectives of multiple stakeholders. This approach was informed by systems thinking tools to understand how interventions may impact a complex, interconnected system. We looked particularly at Causal Loop Analysis - an approach that has been effectively applied in diverse health contexts to uncover underlying dynamics governing service delivery and outcomes, including CVD care models for refugees [24,25]. However, due to worsening security throughout the study period, it was not possible to conduct a confirmatory workshop to finalize the causal loop diagram with participant stakeholders. In spite of this limitation, the authors were able to use the diagram development process and thematic analysis to demonstrate VHWs as critical leverage points for improving cardiovascular outcomes, informing interventions aimed at strengthening community-based support for CVD care delivery and medication adherence [26].

VHWs are a cadre of health workers in the community health system who are from the village where they work. They receive a brief (2-4 week) foundation training and a modest monthly stipend; depending on the specific health project responsibilities and workload, VHWs may receive additional training and payments. Most VHWs have additional responsibilities and sources of income outside the health system [14].

The team developed a semi-structured interview guide to explore factors influencing CVD care and the delivery of NCD and CVD care within the broader healthcare system, and to inform the design of new CVD program activities with VHWs. The guide drew on PP’s prior experience with co-creation of a causal loop diagram for CVD care of refugees in Jordan [25], as well as the health behavior framework and the chronic care model [27,28]. The premise of the health behavior framework is that in order to influence health behaviors that are complex and multi-dimensional, such as participation in screening for CVD risk factors, it is necessary to design interventions that address multiple drivers of the behavior at patient-, provider-, health system, and financing levels. The chronic care model is an effective framework for multi-component care delivery models aimed at improving care processes and outcomes while reducing costs for chronic conditions, including diabetes and hypertension [29].

Participants were selected to include multiple perspectives from a diverse range of stakeholders and roles in CVD care, while balancing the exigencies of an ambitious timeline to transcribe, analyze, and present preliminary findings in the participatory workshop. The sampling strategy aimed to recruit 4 patients, 4 VHWs, 7 EHO clinic staff, and 8 leaders, 8 donors, and 8 NCD experts from different non-governmental organizations (NGOs). Preference was given to individuals with direct experience or expertise relevant to CVD care and the community health system [26,30,31] (See Table). Patient and VHW perspectives were particularly valued in identifying existing limitations of programming and how plans might address identified gaps.

**Table.**
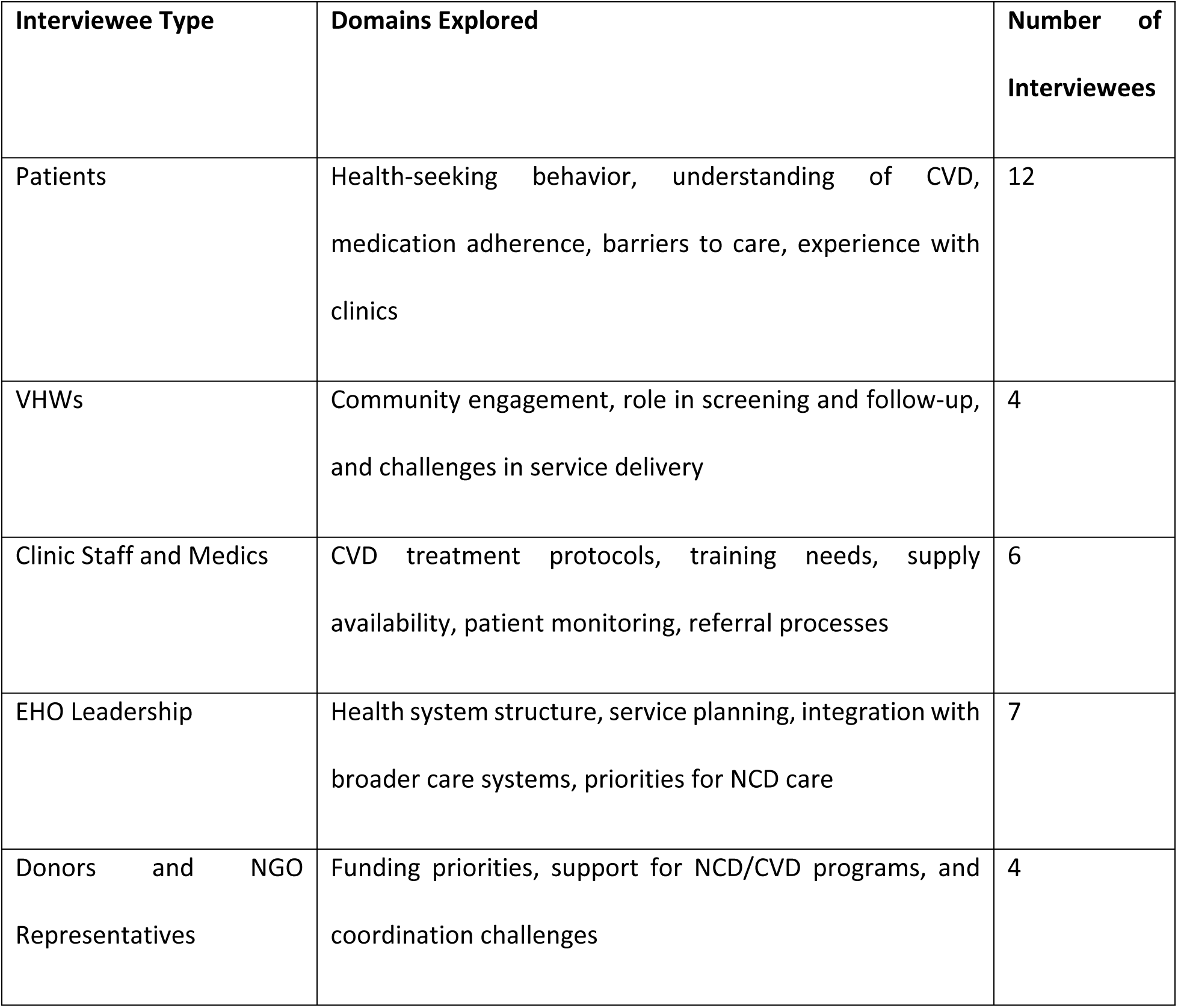
Individuals Interviewed and Domains of Semi-Structured Interviews.

| Interviewee Type | Domains Explored | Number of Interviewees |
| --- | --- | --- |
| Patients | Health-seeking behavior, understanding of CVD, medication adherence, barriers to care, experience with clinics | 12 |
| VHWs | Community engagement, role in screening and follow-up, and challenges in service delivery | 4 |
| Clinic Staff and Medics | CVD treatment protocols, training needs, supply availability, patient monitoring, referral processes | 6 |
| EHO Leadership | Health system structure, service planning, integration with broader care systems, priorities for NCD care | 7 |
| Donors and NGO Representatives | Funding priorities, support for NCD/CVD programs, and coordination challenges | 4 |

All interviews were carried out in the respondent’s preferred language (English, Burmese, or Karen) and both in-person and virtual interviews across Thailand and Eastern Myanmar according to their availability. All key informant interviews (KIIs) provided informed verbal consent, and interviews were audio-recorded, then transcribed and translated into English for analysis [30,31]. The recorded files and transcripts were kept secure and only accessible by study team members. In order to assure safety in an atmosphere of heightened security tensions, names were not recorded. For clarity, roles were noted on each transcript (i.e., EHO leadership, VHW, etc.). This study was deemed exempt by the University of Southern California Office of Human Subjects Research and received ethical approval from the regional Community Ethics Advisory Board, which is a community-led group that provides ethical guidance and oversight for research. Funding was provided by Research for Health in Humanitarian Crises (R2HC/Elrha #78698) and a Research Innovation Award from the Office of Research Excellence, Milken Institute School of Public Health, George Washington University (#09RIA042022).

A first-stage rapid analysis was conducted by PP, with a starting framework of the health system, while being open to new concepts and themes specific to Eastern Myanmar [31]. This initial analysis had to be carried out under time constraints due to security and other field realities. Based on this first analysis, the themes were then organized by the investigators into a preliminary diagram of the health system as it relates to care for CVD risk. This initial analysis informed a written policy brief that was shared with a broad group of stakeholders. Following this, the study team conducted a more formal and in-depth content analysis of the KII transcripts to refine preliminary results, guided by established qualitative content analysis approaches [31]. Two researchers coded the data manually using an Excel matrix. Where there were discrepancies between these code sets, the two coders reviewed the data excerpts and developed a consensus on the most appropriate interpretation. The diagram was revised and reformatted in Canva [32], presenting the identified themes within the health systems as enablers or challenges (or both) in advancing CVD prevention and management in this context.

## Results

We conducted 33 KIIs. The following section describes the relationships, health behavior, health seeking, access, and challenges in CVD delivery at each level of the community health system.

The following illustrative diagram (Fig 1) summarizes the themes, identifying those that are challenges or enabling factors affecting the implementation of VHW-led care for individuals living with CVD across patient, provider, health system, and donor levels.

**Fig 1:**
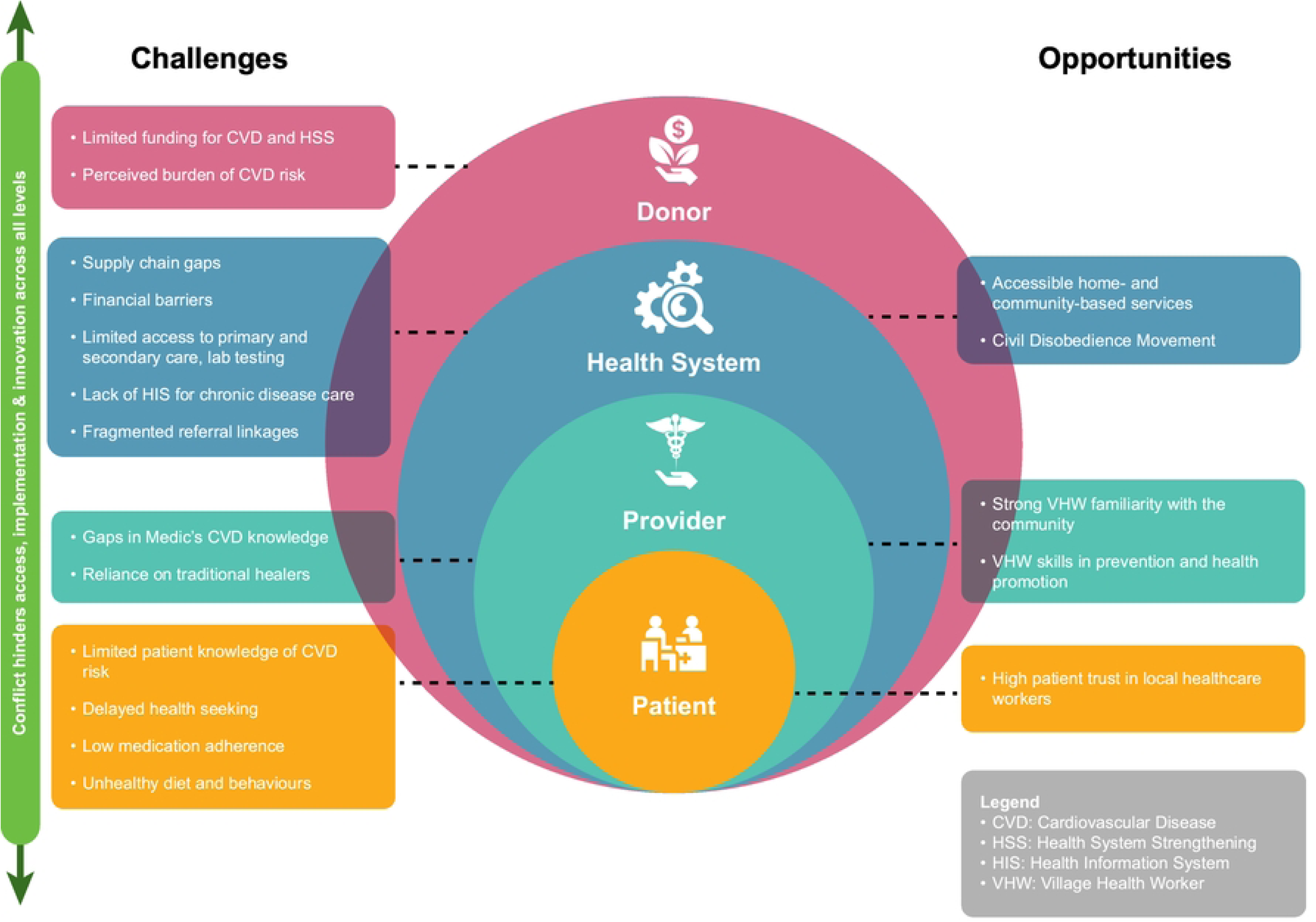
Challenges and opportunities for VHW care models to reduce CVD risk in areas of armed conflict.

Across all levels of care, *conflict* intensifies challenges. Respondents described the conflict leading to clinic closures, staff and patients fleeing for their safety, and transportation became increasingly risky. Security threats, checkpoints, and rising costs disrupted service delivery, supply chains, and affected the accessibility of care, especially for those patients needing referral or chronic disease care.

### Patient Level

#### High patient trust in local healthcare workers

In EHO clinics and villages, trust in local healthcare workers was widespread across Karen communities. Patients mentioned that they sought care at these clinics due to positive experiences and their confidence in the staff. However, when treatment effects were not immediately noticeable, some patients reverted to traditional healers and medicines.

*“I believe that their treatments and quality of care are good, although I am not entirely sure. I simply take the medication they prescribe. They treat the patients nicely.” CVD Patient*

#### Limited patient knowledge of CVD risk

Many individuals lacked basic awareness of CVD and its long-term complications. VHWs and clinic staff mentioned that sharing information among community members could help to spread knowledge about CVD risks and management.

*“Because of the outreach activity, because of health education and community engagement activities, they start to hear about the diseases, and they know they have got the information and also from one person to another person that they will share the information.” EHO Staff*

Strategies such as interactive education sessions, videos, and social group counseling were proposed to enhance patient engagement. Education about the consequences of untreated hypertension and diabetes was also perceived as important to promote medication adherence and manage chronic diseases effectively.

#### Delayed health seeking

Timely health seeking was impacted by each patient’s perception of CVD. The perception that CVD was less critical than other endemic diseases made patients less likely to prioritize CVD care, particularly when they faced other concerns such as financial hardship, safety threats, or transportation challenges. Since initial symptoms are often asymptomatic, patients did not consider CVD as an urgent or serious threat, delaying care-seeking. An interviewed EHO program officer described this as “*They do not care because those diseases may not harm them for the first few years.”.* Moreover, patient adherence to medical advice was also influenced by their perceptions of CVD and constrained by logistical, financial, and educational barriers.

#### Low medication adherence

Some KII respondents were concerned about potential problems with medication adherence. Clinic staff report that many patients stop taking medication once they feel better, and only a few patients follow their treatment consistently. One VHW also stated, “*They [the patients] only go back to the clinics when the diseases reappear again”.* Adherence could be further affected by forgetfulness, cost, lack of understanding, and concurrent illnesses. Moreover, patient adherence to medical advice was also influenced by their perceptions of CVD and constrained by logistical, financial, and educational barriers.

#### Unhealthy diet and behaviors

Although health workers routinely provided diet and behavior advice, they reported that limited local food diversity and difficult living conditions hampered lifestyle changes. Smoking, alcohol, and betel nut use remained common. One program administrator described this:

*“I think the main factor is their stressful living conditions. They have been living in conflict zones along the border since the country’s independence in 1948 (…) This has resulted in a substandard lifestyle where fish paste is consumed more than protein-rich meat.” NGO health program administrator*

### Provider Level

#### VHW familiarity with the community

Participants felt that VHWs who are recruited from communities have unique knowledge and close relationships with the communities they serve. VHWs can serve as a bridge between clinics and remote populations, catalyzing initial engagement with the health system and sustaining continuity of care.

“*VHWs can help organize them without having any social barriers. They can also help in collecting the up-to-date patient’s information while we are away, and can inform us if the patient is tending to not following the medications.” NGO health program administrator*

#### VHW skills in prevention and health promotion

Moreover, VHW skills and knowledge were important for effective chronic disease management. Respondents suggested that VHWs could provide essential services such as home visits, blood pressure and glucose measurement, and health education. VHW respondents described their existing roles managing patient information, assisting medics, monitoring medication adherence, and organizing village health initiatives. However, VHWs were not typically trained in CVD care, limiting their ability to support management of hypertension and diabetes. VHWs mentioned that basic equipment, such as blood pressure cuffs and glucometers, would enhance their capacity to monitor patients and provide timely referrals.

#### Gaps in Medic’s CVD knowledge

Participants described how some medics have extensive experience and training in CVD risk factor management, including lung and heart function, lifestyle changes and prescription of appropriate medications. Many had been trained on protocols from the Burma Border Guidelines, Maternal and Child Health, or Integrated Management of Childhood Illness guidelines. However, some clinical guidelines for CVD was incomplete or outdated, and many providers lack opportunities to practice knowledge gained from didactic trainings in a mentored and supervised visit with real patients. Clinic staff frequently mentioned that both medics and VHWs require updated, focused training to provide evidence-based care to reduce risk.

#### Reliance on traditional healers

Traditional healers continue to play a prominent role in service areas. Several interviewees reported that some patients turn to traditional healers or herbal remedies, particularly when they feel that modern medicine has not improved their condition. One EHO leader also suggested that integrating traditional healers, including traditional birth attendants, into clinic structures may help strengthen community trust and engagement.

*“Maybe we should even include the TBAs in our clinic’s structure to be able to improve our community-based health system. Sometimes we cannot stay at our clinics but have to go to the communities to reach more people.” EHO leader*

*“I took modern medicine, but did not feel better. So, I took traditional medicine and medical plants and roots. After taking them, I got better.” CVD Patient*

### Health System Level

*“The military coup has had a significant impact on healthcare in the region, leading to delays in the transportation of medical supplies, difficulties in accessing advanced healthcare facilities in towns, and security concerns due to the presence of multiple checkpoints.” EHO Clinic Staff*

*“When they get information that the airstrike is coming this way or to this area, the other junta soldiers are burning the house, [the health workers] have to run(…)In this situation, we [as their leaders] can’t do anything. We can’t blame them “Why aren’t you treating the patient? Why aren’t you measuring blood pressure?”(…)it’s affecting providing medical services and I think there’s no answer for that.” EHO coordinator*

#### Supply chain gaps

Availability and continuity of medicine supplies were major challenges across clinics, reported by both health workers and patients. The lack of blood pressure cuffs and glucometers limited providers’ ability to conduct proper diagnoses. Medication stockouts were common, leading some clinics to instruct patients to buy from local pharmacies. Therefore, maintaining continued access to medications was a significant concern for both providers and patients. When patients are diagnosed with a chronic disease, they are initially prescribed monthly dosages, which is extended to two-three months as symptoms stabilize. Giving medications for two to three months is more convenient for both providers and patients, but careful monitoring is needed to ensure patient follow-up and adherence monitoring.

*“The clinic staff initially prescribed a monthly dosage. However, now that they trust me to take my medicines regularly, I am receiving a three-month dosage. They always check if I have any medicine left from the previous dosage.” CVD Patient*

#### Financial barriers

*“Inflation since the military coup has made it difficult for many of us to afford the medications we need. I could not afford it if I had to buy the monthly medicines I have to take.” CVD Patient*

Medications are ostensibly available free of charge from KDHW clinics, but some CVD patients seek care from other providers, and several stakeholders described substantial financial barriers to care and medication. Many patients could not afford to complete full courses of treatment, often relying on sharing prescriptions or buying small amounts at a time. While the cost of diagnosis and consultation is generally fee-free for most EHO clinics, some patients may donate or pay a nominal fee. While this is not expected from all patients, some respondents believed this could be a barrier for some low-income patients. For acute and complex cases, the cost of transportation to higher-level care, particularly for cross-border referrals to Thai clinics, could be prohibitively expensive. Some, but not all, Thailand-based clinics provide limited fee-free secondary care to alleviate this.

#### Accessible home- and community-based services

Respondents believed village-level services for screening (e.g., screening among the community for diabetes, hypertension, obesity, or smoking habits), home-based treatment follow-up, and linking them to appropriate care facilities were effective strategies. Some respondents suggested that mobile outreach activities could be beneficial, but these require trained volunteers and strong monitoring systems.

#### Access barriers to primary care for CVD risk

One of the major challenges for clinics to manage CVD was a lack of trained professionals, with staff not trained for complicated cases. Some EHO leaders pointed out challenges such as long distances, high transportation costs, and low awareness of services, which continued to limit access and discourage clinic attendance.

#### Access barriers to secondary care

Guidelines recommend that complicated cases with signs and symptoms of acute stroke or heart attack be referred to hospitals for confirmation of diagnosis and specialized treatment. Travelling to these facilities sometimes involved crossing the border into Thailand. Transportation costs for some referrals might be covered based on the patient’s symptoms and the availability of facilities at the referral center; however, coverage of such costs is conditional on clinic cash reserves, donor support, and often depends on time- or geographically-limited projects.

“*Some patients…can afford to pay the transportation charges themselves. For those who could not pay for it, we pay the cost with the clinic’s funds for patient referrals. I think there is a limit for the transportation expenses per patient.*” EHO clinic staff

#### Civil Disobedience Movement (CDM)

During KIIs with the EHO leaders and donors, they stated that healthcare workers who fled government crackdowns have helped to fill gaps in secondary care since the coup. CDM professionals contribute to mobile medical units, provide training, and identify CVD risk factors in the community. Nonetheless, ongoing, locally based continuous assistance, including medicine availability, is required to enhance their effectiveness, and it is not clear how long recently arrived clinicians, many of whom previously lived in urban centers of central Myanmar, will remain in remote rural border areas.

#### Limited access to laboratory testing

While some clinics have basic diagnostic tools like malaria microscopy, many clinics experienced what one EHO staff member described as *“numerous insufficiencies”* in basic equipment and capacity. Patients need to travel to the nearby city if they need even basic laboratory tests (e.g., serum creatinine or lipid profile). Supply chain challenges and equipment shortages hinder effective diagnostics. Some EHO leaders from the KIIs suggested expanding access to diagnostic tools and offering more frequent training to strengthen capacity.

#### Lack of HIS for chronic disease care

Medics recorded clinical information using paper records and Excel spreadsheets that made it difficult, if not impossible, to view information from prior visits. Monthly reports were submitted online to the headquarters. However, some clinic staff stated that due to the current political situation, data and statistics were not up to date. Data entry errors and a lack of standardized guidelines for follow-up tracking caused challenges in decision-making and program planning for leadership.

#### Fragmented referral linkages

Referral processes were fragmented and increasingly difficult after the coup. According to KIIs with patients, they faced language barriers, safety risks, rising costs, and travel restrictions. These restrictions forced changes in the referral process, and patients were reluctant to travel. Some clinics stopped providing referral letters due to patient safety concerns. The lack of clear EHO referral protocols also impacted the quality and efficiency of referrals. Suggestions from staff and patients included improving referral systems, increasing staff capacity, ensuring better coordination with other providers, and developing clinic-based systems for tracking follow-up and medication adherence.

### Donor Level

#### Limited funding for CVD risk factor management

Participants reported that while the burden of CVD has risen, the resources allocated to addressing it remain very low. According to EHO leaders, CVD services largely relied on referral to other service providers in Thailand. Most funding to Eastern Myanmar continued to prioritize endemic communicable diseases, which were perceived as more urgent and locally manageable. Chronic conditions like CVD, which require long-term management and investment, received comparatively less attention, especially in active conflict settings.

*“ We were very busy with the COVID response and a [then the] coup happened(….)we try to shift focus more on this emergency response(…)so, people are very busy and overloaded with these ongoing conflicts and also system strengthening focus. And also, I think [this is] also the donor’s perspective. So, the NCD was overlooked.” Donor*

#### Shifts in funding for primary care health system strengthening (HSS)

HSS projects have gained donors’ interest, particularly around improving basic health service deliveries and governance among local communities. However, this funding focused on the current service packages (communicable diseases and maternal and child health). Moreover, the ongoing crisis shifted donors’ priorities toward emergency humanitarian support. EHOs also faced structural challenges in accessing funds, including the absence of a unified health information system, which contributed to difficulties in monitoring and reporting.

The community health system is dependent on international funding. Modest local financing exists, but individuals have very little capacity to pay. The lack of local payments at clinics reduces the burden on patients, but means that clinics have no consistent local funding. The lack of a defined tax base or revenue model in local areas raised concerns about the sustainability of healthcare financing. Even though some clinics benefited from volunteer and CDM doctors, these opportunities were fragile and required additional investment in supplies, medications, and capacity building to be sustained.

#### Ignorance of the high CVD burden

Although concern about CVD was increasing locally, the donors’ perception was that it took time to see outcomes and was difficult to monitor. Donors suggested that to receive more support for NCD care, it’s important to raise awareness using data and advocacy, and by linking CVD efforts with broader health system improvements.

## Discussion

This study describes qualitative findings from stakeholder interviews to inform development of the first village health worker-led intervention to improve screening and access to care for CVD in a conflict-affected region of Myanmar. Previous studies have described development of care models for refugee populations in relative stable countries such as Jordan and Lebanon [25,33]. Our study extends this work to explore possible roles for VHWs in active conflict zones [4,5,34]. In the context of this project, we have consulted with individuals with a stake in CVD care in an area of Eastern Myanmar with protracted and escalating conflict. Our final analysis and care model highlight local strengths and opportunities of taking a VHW-led approach.

Findings presented here contributed to a workshop to develop a VHW care model that involved universal screening of adults 40 years and older for CVD risk factors, and enrollment of eligible villagers in monthly household visits by the VHW. This model subsequently was shown to be acceptable and feasible to implement in three villages [35]; a cluster randomized controlled trial is testing the impact of the care model on medication adherence, CVD risk and cost effectiveness [35].

Based on the results of the analysis, the authors identified five areas of influence for VHWs, as well as associated challenges in this context. The first four have relevance to both conflict and stable settings, however the acute conflict setting brings specific roles for VHWs. The fifth area of influence describes the completely unique role that VHWs have in active conflict zones, present and responsive through all changes in the conflict and in community circumstances.

### 1. VHW-Led Community-Based Prevention via Risk Factor Modification

This study suggests that VHWs are uniquely positioned to deliver culturally tailored interventions that promote risk factor modification, an approach supported by multiple studies highlighting the effectiveness of VHWs in NCD prevention [15,25]. Low awareness of CVD among patients and health workers was a critical barrier according to the study findings. Respondents agreed that VHWs might fill this awareness gap by conducting regular health education sessions, promoting smoking cessation, supporting healthy diet and physical exercises, and emphasizing on early screening and continued medical adherence.

Transportation barriers and the current ongoing conflict limit the ability of villagers to access clinic-based care, underscoring the importance of VHW-led outreach. Through home visits and community education sessions, our study respondents believed that VHWs can raise awareness of long-term CVD risks, emphasize preventive measures, and link community members to clinic resources. This multifaceted approach has proven effective in low-resource settings where healthcare systems function as complex adaptive entities requiring locally driven strategies [15,36,37], and respondents feel that VHWs can play similar roles in war-torn areas

### 2. VHW Screening of Communities for Elevated CVD Risk, Diabetes, and Hypertension

Many community members only seek care when symptoms become severe, often missing the window for early intervention. A study in rural Uganda demonstrated that VHWs can fill this gap by conducting blood pressure and diabetes screening and monitoring in the community, reducing the need for travel to distant clinics [38]. However, there is a global debate on the value of community-based screening. For example, critics of community screening for hypertension note that some programs suffer from inaccurate blood pressure assessment, low rates of follow-up, and treatment adherence [39]. Our VHW-led CVD care model screening approach will need to ensure that the VHW then actively follows up with the individual, and that perhaps support from a trusted VHW from the community can, in fact, encourage behavior change.

In the current community health system, where travel is unsafe or costly, VHWs serve as the initial contact, identifying at-risk individuals and facilitating referrals for endemic infectious diseases. As in other contexts, ongoing training and capacity-building will be crucial to making sure VHWs can perform accurate screenings, interpret data correctly, and educate patients on risk factor management [40–42].

### 3. VHW Support for Adherence to Medication, Disease Management, and Clinical Care

Low medication adherence came out as an important concern, with patients often discontinuing treatment once they feel better, or due to financial constraints, or due to a lack of understanding of the chronic nature of CVD. Costs for accessing care and medicine supply chain challenges are deepened by political instability and displacement [43]. Moreover, when clinics close or staff relocate due to active conflict, patients may lose contact with their regular healthcare providers. In these unstable conditions, some respondents feared that incorrect information on CVD and treatment spreads more easily, which may reinforce poor adherence and delayed health seeking.

According to stakeholders, with additional basic training VHWs can also provide monthly monitoring of blood pressure and (when applicable) blood glucose. By building on to the existing reporting pathways, they can then report their findings to the regional medic on a monthly basis for review. This emphasizes the importance of frontline agents (like VHWs) to improve the reach and timeliness of care in conflict-affected regions [41].

Study respondents in supervisory and leadership roles believe that VHWs can provide vital support for adherence via home visits and consistent follow-up by reminding patients about dosing, clarifying side effects, and reinforcing the importance of continuous medication, even when symptoms subside. Previous research has shown that VHW interventions can improve medication adherence for chronic conditions in low and middle-income countries [15,18,35,37,44]. Moreover, VHWs can quickly relay shortages or clinical concerns back to EHOs, ensuring that gaps in healthcare are addressed.

### 4. Standardized Referral Pathways (From Clinic) and Follow-Up (To Clinic)

The study highlights major gaps in the referral system for CVD patients in Eastern Myanmar. Providers reported that financial constraints, transportation difficulties, and security risks limit access to advanced care. Moreover, the lack of standardized referral guidelines and follow-up mechanisms makes patient referrals challenging. Border restrictions, high costs, and poor coordination hinder access, similar to other studies [25,43]. Even when referrals occur, tracking patient outcomes is still difficult.

To improve this, experienced providers in our KII suggested a standardized referral system could set clear criteria for CVD cases, safe referral pathways, and systematic tracking for follow-ups. Strengthening VHW roles in post-referral support through home visits and digital health tracking could improve adherence and ensure better long-term management [35,45].

### 5. Responsive to an evolving context

Subsequent to the period of data collection for this study, additional challenges to chronic disease care in Eastern Myanmar have emerged, which we acknowledge here and will inform future planning. The conflict in Southeastern Myanmar has evolved, with increasing air strikes in the region. Myanmar experiences some of the highest levels of attacks on healthcare workers and facilities [46]. Arguably, the role of VHWs, who may relocate with their fellow villagers when forced to relocate, remains more essential than ever. Despite many challenges, such as limited resources, insecure environments, and shifting donor priorities toward humanitarian and communicable disease programs, VHWs can fill the gaps in screening, medication adherence, and psychosocial support. VHWs also provide a low-cost means of extending clinical care in a shrinking funding environment.

With the worsening insecurity, VHWs may also have a role in supporting patients to manage mental health symptoms. Conflict-affected populations face chronic stressors such as displacement, ongoing violence, and economic hardship, which can worsen both mental health conditions and physical health, like hypertension or diabetes. Several respondents in our study mentioned that fear, stress, and financial worries made it harder for patients to manage their health and continue taking medicines. Clinic Staff also mentioned that they had concerns with medication adherence due to these unstable conditions. Traditional beliefs and stigma may discourage patients from seeking help, further complicating CVD management [47,48]. Training VHWs in psychosocial support can help address these stressors at the community level, and can improve treatment adherence and outcomes [49], and promote a more holistic approach to healthcare [49,50].

## Limitations of the study

Patients, VHWs, and other clinical staff interviewed for this study were residents in remote rural areas of Eastern Myanmar served by a single EHO, which may limit generalizability to other areas of Myanmar or to armed conflicts in other regions.

Some community members and VHWs may not have felt comfortable engaging fully, cautious of topics perceived to be sensitive to funding decisions, or a more general need to be circumspect when discussing the details of service delivery in an active conflict zone. Communication and transportation barriers, worsened by active conflict, made several interviews challenging.

Due to worsening security during this period, it was not possible to have a final workshop to confirm with stakeholders the complex care system represented by the draft causal loop diagram. In spite of this limitation, the authors were able to use the draft diagram and thematic analysis to demonstrate that the strengths of VHWs can be leveraged to improve cardiovascular outcomes, informing interventions aimed at strengthening community-based support for CVD care adherence and delivery [26,30].

## Conclusion

This qualitative study underscores the urgency to develop and test VHW care models to mitigate the growing burden of CVD in conflict-affected Eastern Myanmar. Study respondents endorsed the capacity of VHWs to play multiple constructive roles in CVD management for populations confronting dual burdens of chronic disease and conflict. Our study may hold implications for other academic-agency research partnerships interested in employing similar qualitative methods to inform the co-creation of care models designed to reduce CVD risk in acute crises, such as the VHW care model being tested in a cluster randomized controlled trial in Eastern Myanmar.

## Data Availability

The data that supports the findings of this study is available upon reasonable request from the corresponding author, AR.

## Acknowledgments

We thank all key informant interview participants, including community members, VHWs, clinic staff, EHO leaders, and donor representatives, for generously sharing their time and insights. We particularly appreciated the data collection team’s dedication and the leadership in Karen State for their support throughout the study. In particular, we would like to specifically acknowledge Saw Diamond Khin, the Executive Director from the Karen Department of Health and Welfare, for his continuous collaboration and invaluable contributions to the study.

